# COVID-19 infection induces readily detectable morphological and inflammation-related phenotypic changes in peripheral blood monocytes, the severity of which correlate with patient outcome

**DOI:** 10.1101/2020.03.24.20042655

**Authors:** Dan Zhang, Rui Guo, Lei Lei, Hongjuan Liu, Yawen Wang, Yili Wang, Hongbo Qian, Tongxin Dai, Tianxiao Zhang, Yanjun Lai, Jingya Wang, Zhiqiang Liu, Tianyan Chen, Aili He, Michael O’Dwyer, Jinsong Hu

**Affiliations:** Department of Cell Biology and Genetics, Xi’an Jiaotong University Health Science Center, Xi’an, China; Clinical Laboratory, Xi’an No.8 Hospital (Shaanxi Infectious Diseases Hospital), Xi’an, China; Department of Critical Care Medicine, The First Affiliated Hospital, Xi’an Jiaotong University, Xi’an, China; Biobank, The First Affiliated Hospital, Xi’an Jiaotong University, Xi’an, China; Department of Pathology, Xi’an Jiaotong University Health Science Center, Xi’an, China; Department of Epidemiology and Biostatistics, School of Public Health, Xi’an Jiaotong University Health Science Center, Xi’an, China; Clinical Laboratory, Xi’an No.9 Hospital, Xi’an, China; Tianjin Key Laboratory of Cellular Homeostasis and Human Diseases, Department of Physiology and Pathophysiology, School of Basic Medical Science, Tianjin Medical University, Tianjin, China; Department of Infectious Diseases, The First Affiliated Hospital, Xi’an Jiaotong University, Xi’an, China; Department of Clinical Hematology, The Second Affiliated Hospital, Xi’an Jiaotong University, Xi’an, China; Apoptosis Research Centre, Biomedical Sciences, National University of Ireland Galway, Ireland; The Institute of Infection and Immunity, Xi’an Jiaotong University Health Science Center, Xi’an, China

## Abstract

**Background:** Excessive monocyte/macrophage activation with the development of a cytokine storm and subsequent acute lung injury, leading to acute respiratory distress syndrome (ARDS) is a feared consequence of infection with COVID-19. The ability to recognize and potentially intervene early in those patients at greatest risk of developing this complication could be of great clinical utility.

**Methods:** We performed detailed flow cytometric analysis of peripheral blood samples from 28 COVID-19 patients treated at Xian No.8 Hospital and the First Affiliated Hospital of Xian Jiaotong University in early 2020 in an attempt to identify factors that could help predict severity of disease and patient outcome.

**Findings:** While we did not detect significant differences in the number of monocytes between patients with COVID-19 and normal healthy individuals,we did identify significant morphological and functional differences, which are more pronounced in patients requiring prolonged hospitalization and ICU admission. Patients with COVID-19 have larger than normal monocytes, easily identified on forward scatter, side scatter analysis by routine flow cytometry,with the presence of a distinct population of monocytes with high forward scatter (FSC-high). On more detailed analysis, these FSC-high monocytes are CD11b^+^, CD14^+^, CD16^+^, CD68^+^, CD80^+^, CD163^+^, CD206^+^ and secrete IL-6, IL-10 and TNF-alpha, consistent with an inflammatory phenotype.

**Conclusions:** The detection and serial monitoring of this subset of inflammatory monocytes using flow cytometry could be of great help in guiding the prognostication and treatment of patients with COVID-19 and merits further evaluation.

## Introduction

Severe Acute Respiratory Syndrome coronavirus 2 (SARS-CoV-2), also known as Corona Virus Disease-19 (COVID-19) is a new coronavirus, first identified in Wuhan, China in December 2019, which frequently induces fatal inflammatory responses and acute lung injury.^1,2^ As of March 20 2020 a total of 245,484 confirmed cases have been reported worldwide, with an estimated mortality of 4% (https://coronavirus.jhu.edu/map.html).

Patients experience a spectrum of disease from a mild illness, through varying severity of pneumonia all the way to ARDS and sepsis with multi-organ failure and death. Initial clinical features at disease onset are fever (77-98%), dry cough (46-82%), myalgia or fatigue (11-52%) and dyspnea (3-31%).^3^ The majority of patients develop pneumonia, which can proceed in up to 20-30% of cases to respiratory failure requiring intubation and ventilatory support. In those COVID-19 patients who go on to develop pneumonia, dyspnea develops a median of 8 days after onset of illness. Radiographic abnormalities (CT or chest x-ray) consisting of ground-glass opacities and focal consolidation are seen in patients with pneumonia. Major causes of death include respiratory failure and myocardial damage due to myocarditis. Acute kidney injury, secondary infection and coagulopathy are each seen in approximately 50% of non-survivors. Mortality increases with age and in patients with underlying co-morbidities, such as hypertension, diabetes mellitus, coronary heart disease, chronic lung disease, and cancer. According to a recent retrospective report from Wuhan, clinical predictors of increased mortality on multivariate analysis included advanced age, progressive organ failure and elevated D-Dimers on admission.^4^ Other factors significantly associated with poor outcome on univariate analysis included elevated levels of serum ferritin, interleukin-6 (IL-6), Alanine amino transferase (ALT), lactate dehydrogenase (LDH) and highly sensitive cardiac Troponin I, as well as reduced levels of lymphocytes, Hemoglobin (Hb), Platelets and serum albumin.

The best recognized hematologic abnormality is lymphopenia, which is seen in up to 85% of severe cases with the severity of lymphopenia linked to outcome.^5^ Since regulatory T cells play an important role in dampening the immune response, excessive elimination of these cells could result in an unchecked innate immune responses, leading to an uncontrolled inflammatory response.^5^ Indeed, there is growing evidence implicating excessive monocyte/macrophage activation and associated cytokine storm with the pathophysiology of severe SARS-CoV-2 disease related complications.^6^ Despite this, there are few reports to date relating to abnormalities of monocytes in patients with COVID-19. Herein we describe novel observations in relation to changes in monocyte morphology and activation status, which correlate with the prognosis and severity of COVID-19 infection and which can be readily quantified by flow cytometry with the concurrent measurement of forward scatter (FSC) and (SSC), which measure cell size and complexity, respectively. Specifically, we have identified the presence of a population of monocytes with higher FSC than normal (FSC-high) not typically seen in other types of viral infection. Further analysis reveals that these FSC-high monocytes express CD14, CD11b, as well as CD16 along with markers associated with both M1 and M2 polarization, and secrete IL-6, IL-10 and TNF-alpha.

## Methods

### Patients

In this study, 28 cases of COVID-19 were from Xi’an No.8 Hospital (Shaanxi Provincial Infectious Disease Hospital) and the First Affiliated Hospital of Xi’an Jiaotong University, which are the designated hospital for COVID-19 by the local government (Shaanxi province, China). All COVID-19 patients were diagnosed according to the World Health Organization (WHO) interim guidance (https://www.who.int/publications-detail/infection-prevention-and-control-during-health-care-when-novel-coronavirus-(ncov)-infection-is-suspected-20200125) and the Guide of Diagnosis and Treatment of COVID-19 (6th edition, in Chinese) published by the National Health Commission of China (http://www.nhc.gov.cn/yzygj/s7652m/202002/54e1ad5c2aac45c19eb541799bf637e9.shtml), and confirmed by SARS-CoV-2 nucleic acid testing of throat swab specimens using RT-PCR (Realtime PCR). This study was approved by the Ethics Commissions of Xi’an No.8 Hospital and the First Affiliated Hospital of Xi’an Jiaotong University (2020-07), with a waiver of informed consent due to a public health outbreak investigation. The study was registered at http://114.255.48.20, which is the Medical Research Registration Information System of the National Health Commission of the People’s Republic of China.

### Blood samples

Blood samples from patients were collected for laboratory assessments according to the doctor’s instruction. All patients had not received any treatment before blood sampling.

### Flow cytometry analysis

For membrane staining, 50 μL K3-EDTA anti-coagulant whole blood cells were incubated with a panel of fluorochrome-labeled antibodies or unstained/FMO (fluorescence minus one) controls for 15 min at room temperature. Erythrocytes were further lysed by mixing with 450 μL BD FCAS lysing solution (BD Biosciences, San Jose, CA, USA; Catalog No. 349202) for 5-10 min. After washing cells by adding 2 to 3 mL PBS and centrifuge for 5 min at 400g, the cells were resuspended in 400 μL PBS and examined by a flow cytometer (BD FACScantoTM II, BD Biosciences, San Jose, CA, USA) using the FACSDiva v. 6.1 software. The data were analysed by FlowJo software (Version 7.6.1; Tree Star, Inc., Ashland, OR, USA).

For intracellular staining, aliquot 100 μL fresh whole blood per assay tube. Then, surface marker staining was performed first as mentioned above. After lysis of red blood cells and rinse with PBS, the samples were then fixed with 1% paraformaldehyde in PBS for 15 min, permeabilized with 0.1% saponin for 30 min at room temperature. After rinsing the samples with PBS, the cells were incubated with antibodies for staining intracellular molecules for 30 min. Next, the cells were washed by centrifugation in 3 mL PBS twice. Resuspended cells in 0.4 mL PBS were analyzed on the flow cytometer.

To investigate the expression of the SARS-CoV-2 receptor ACE2 (angiotensin converting enzyme 2) on monocyte/macrophage cell lines, an indirect staining was performed as following. Briefly, human monocytic cell lines THP-1 and U939, as well as murine macrophage cell line RAW264.7 were cultured in RPMI1640 medium supplemented with 10% FBS (fetal bovine serum) and antibiotics, or high glucose DMEM medium supplemented with 10% FBS but without antibiotics (for RAW264.7), at 37□ in a 5% CO_2_ air incubator. Then, half million of log-phase cells were then collected to stain with the primary ACE antibody for 15 min at room temperature. After washing with PBS, the cells were further stained with fluorochrome-labeled second antibody for 15 min. The washed and resuspended cells were finally analyzed on the flow cytometer.

A full list of antibodies used in this study is included as Supplementary File 1.

### Wright’s staining of the blood smear

The peripheral blood smears were made and stained with Wright’s stain using a SP1000i (Sysmex, Kobe, Japan) automated smear-maker-stainer system. Examination of stained blood smears were semiautomated with a digital cell morphology system CellaVision DM96 (CellaVision AB, Lund, Sweden), following the manufacturer’s instructions.

### Survival analysis for discharging of COVID-19 patients from hospitals

To investigate the potential effects of FSC-low monocytes and FSC-low/FSC-high monocytes on the discharge of COVID-19 patients from hospital, we performed Kaplan-Meier survival analysis. 24 COVID-19 patients were categorized into high FSC-low monocytes % and low FSC-low monocytes % groups based on the median of FSC-low monocytes % values. Survival curves were obtained for both groups and log-rank test was implemented to examine the statistical significance of the difference in survival curves for the two groups. Similar analysis was also conducted for FSC-low/FSC-high monocytes. In addition, Cox models were also fitted to adjust the potential effects of age and gender.

### Statistical analysis

Statistics values are presented as means ± SD. Data were analyzed by using GraphPad Prism version 6.04 (GraphPad Software, San Diego, CA, USA). Statistical significance was calculated by Student’s unpaired t-test, where differences at *P* < 0.05 was considered statistically significant.

## Results

### A specific FSC-high population can be identified in the peripheral blood of COVID-19 patients

To better understand the impact on the immune response of SARS-CoV-2/COVID-19, we originally sought to investigate changes of immune cells in the peripheral blood in COVID-19 patients using flow cytometry. Unexpectedly, in all tested COVID-19 patients, we found the presence of a specific population right next to the population of monocytes (FSC-high), when using the FSC and SSC parameters (Figure 1A). In contrast, this population is virtually absent in healthy donors, (Figure 1B-C). To validate the nature of the FSC-high population, we further performed Wright-Giemsa staining on blood smears. As shown in Figure 1D, we confirmed the presence of an increased number of larger, atypical, vacuolated monocytes, not normally seen in the peripheral blood of healthy individuals. Table 1, shows that while the total number of monocytes/macrophages in the peripheral did not differ between patients and a healthy donor, there was an increased proportion of activated monocytes/macrophages in patients with COVID-19.

**Table 1.** Morphological changes of monocytes in peripheral blood of HD and COVID-19.

|  | **The percentage of monocytes and**<br>**macrophages per 200 blood cells** | **The ratio of monocytes/macrophages per 200**<br>**blood cells** |
| --- | --- | --- |
| Healthy donor | 4.2% | 9/0 |
| Patient 1 | 7.9% | 3/12 |
| Patient 2 | 4.6% | 2/5 |
| Patient 3 | 2.1% | 1/3 |
| Patient 4 | 4.0% | 0/8 |

**Fig 1.**
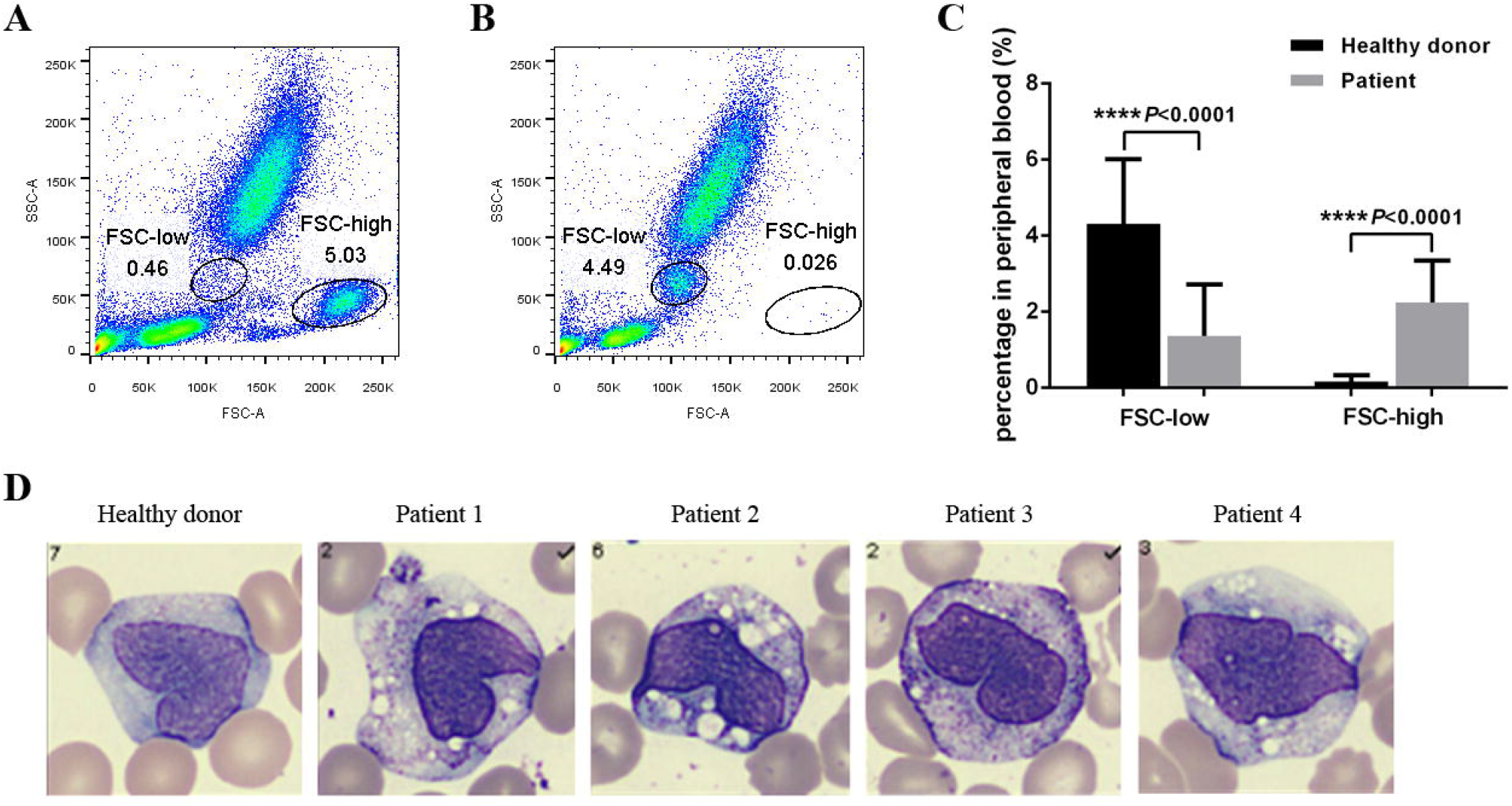
Flow cytometry analysis of the peripheral blood identified a FSC-high population in COVID-19 patients. (A-B) Flow cytometry parameter FSC/SSC identifies a specific population in the peripheral blood of COVID-19 patients. (C) Statistical analysis of the percentage of FSC-low and FSC-high population in healthy controls and COVID-19 patients (HD n=16, patient n=28). (D) Representative pictures of the FSC-high monocytes in COVID-19 (× 1000 magnification).

### The FSC-high population in COVID-19 expresses macrophage markers

Next, we analyzed the expression of the phenotypic markers on the FSC-high population using flow cytometry. As shown in Figure 2A-B, these cells are strongly positive for CD14 and CD16, suggesting that they belong to the monocyte lineage. More importantly, macrophage markers CD11b, CD68, CD80, CD163, CD206 are all expressed in this population. Since CD80 is considered a marker typical of M1 polarization, and CD163 and CD206 are considered to be typical of M2 polarization, it suggests that the FSC-high population contains both M1 and M2 macrophages. To further confirm this suspicion, we performed intracellular staining to validate the expression of M1/M2-specific cytokines in the FSC-high cells. As shown in Figure 2C-D, M1-specific cytokines IL-6 and TNF-α were found to be secreted, while on the other hand, the M2-specific cytokine IL-10 was also expressed by cells in the FSC-high population. Collectively, these findings strongly support that the FSC-high population in COVID-19 are composed of macrophages that have a mixed M1 and M2 polarization.

**Figure 2.**
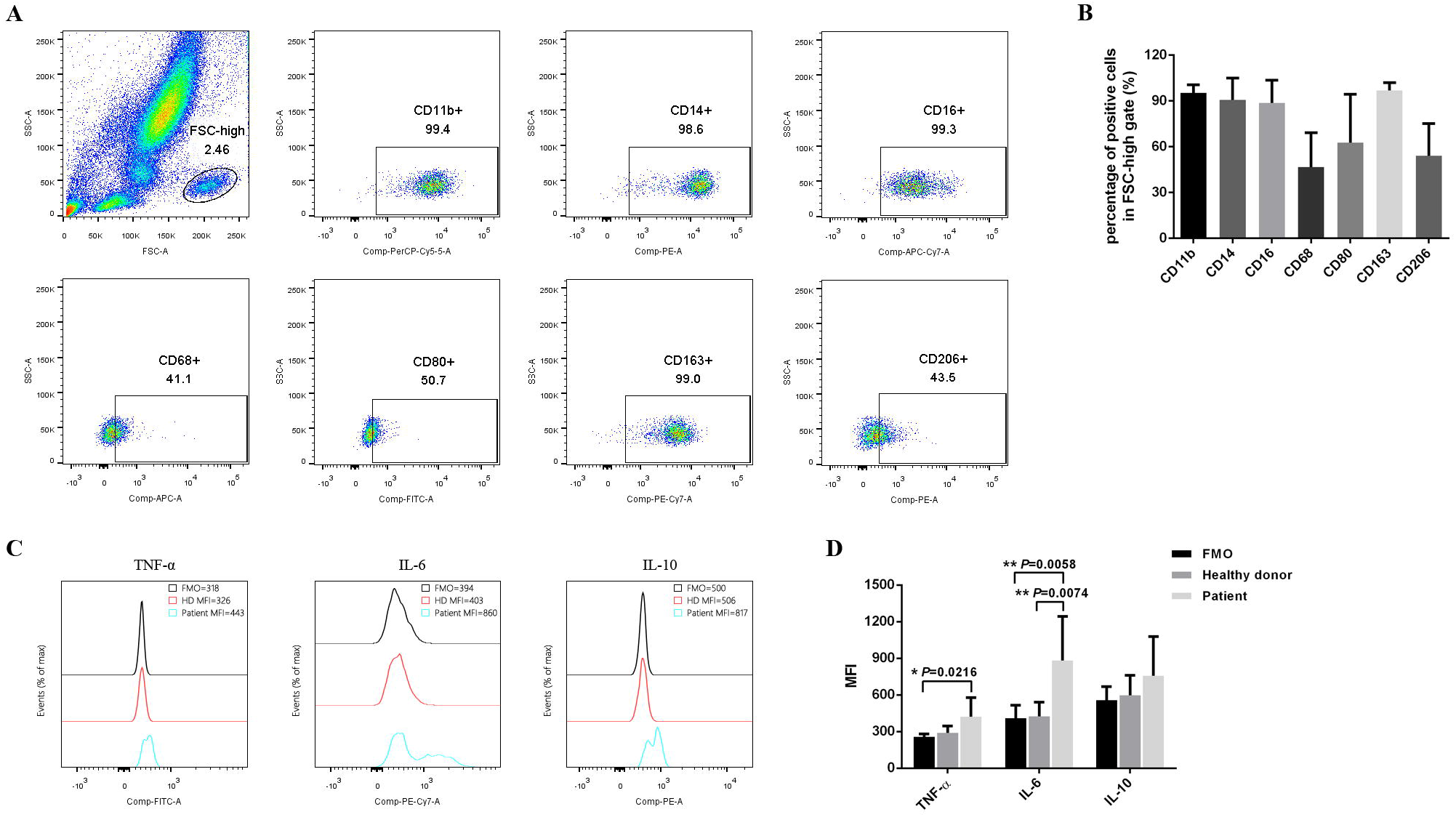
Flow cytometry analysis of the expression of the macrophage markers on the cells in FSC-high population. (A-B) Membrane staining shows the expression of monocyte/macrophage related markers (n=14). (C-D) Intracellular staining analysis of the expression of M1/M2-asssociated cytokines (HD n=6, patient n=15).

### The monocytes/macrophages in COVID-19 contain a decreased number of classical, with an increase in intermediate and nonclassical subsets

Given that macrophages are originally differentiated from monocytes, we further compared the distribution of the classical, intermediate and nonclassical subsets of monocytes in COVID-19 patients and healthy donors by counting the percentage of CD14^++^CD16^-^, CD14^++^CD16^+^, and CD14^+^CD16^++^ cells in FSC-low and FSC-high populations. As shown in Figure 3, in contrast to heathy donors, the number of classical monocytes in COVID-19 patients decreased, but the number of intermediate and nonclassical monocytes increased. This suggests that the monocytes in COVID-19 patients are different from those in healthy donors.

**Figure 3.**
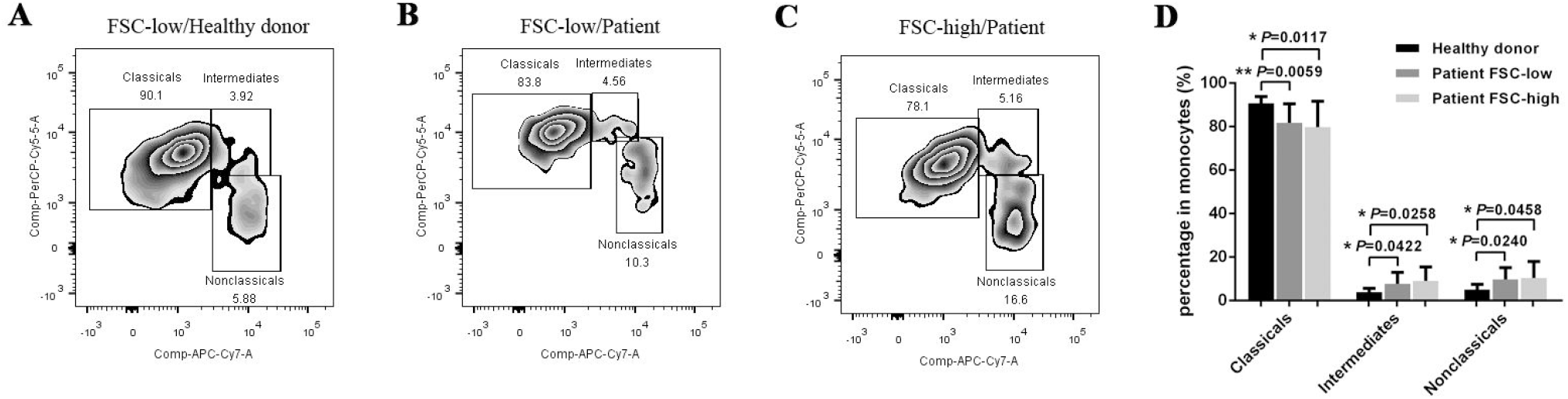
The percentage of the three kinds of monocyte subsets (classical/intermediate/non-classical) in FSC-low and FSC-high populations. (A) The percentage of the classical/intermediate/non-classical monocytes in FSC-low of healthy controls. (B) The percentage of the classical/intermediate/non-classical monocytes in FSC-low of COVID-19 patients. (C) The percentage of the classical/intermediate/non-classical monocytes in FSC-high of COVID-19 patients. (D) Statistic analysis shows that compared to healthy controls patients had a reduction in classical monocytes with a higher proportion of intermediate and non-classical monocytes (HD n=9, patient n=20).

### SARS-CoV and SARS-CoV-2 receptor ACE2 is strongly expressed in monocytes

ACE2 has been shown to be the entry receptor for both the SARS-CoV and SARS-CoV-2.^7-10^ Speculating that direct viral infection could trigger the observed changes in monocytes, we further investigated whether monocytes express ACE2 and therefore could be infected by the SARS-CoV-2. By performing flow cytometry staining ACE2 on human monocytic cell lines THP-1 and U939, as well as murine macrophage cell line RAW264.7, we demonstrated that all these monocyte/macrophage cells are ACE2 positive (Figure 4A). Moreover, we confirmed that the monocytes in the peripheral blood of the healthy donors and COVID-19 patients are positive (Figure 4B-D). More importantly, we found that the expression level of ACE2 on the monocytes/macrophages in COVID-19 patients is significantly lower than healthy donors (Figure 4E).

**Figure 4.**
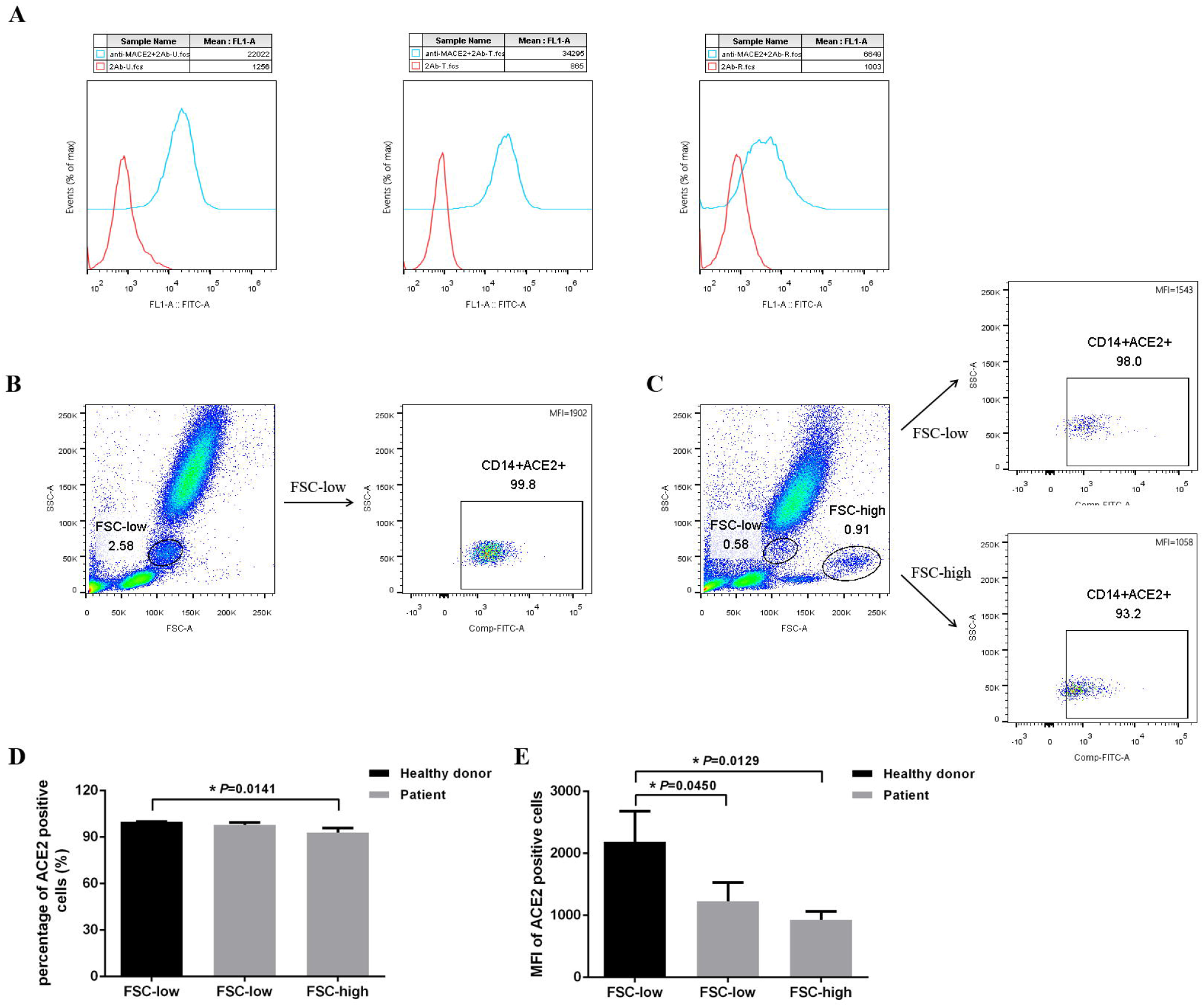
SARS-CoV-2 receptor ACE2 is positively expressed on the surface of the monocytes. (A) Flow cytometry analysis of the expression of ACE2 on monocyte/macrophage cell lines. (B-C) Flow cytometry analysis of the expression of ACE2 on monocyte/macrophage in peripheral blood of the heathy controls and COVID-19 patients. (D-E) Statistical analysis shows that compared to healthy donors, patients had lower expression of ACE2 on their monocytes (HD n=3, patient n=3).

### Survival analysis showed that FSC-low monocytes and FSC-low/FSC-high monocytes were associated with time to discharge from hospital for patients with COVID-19

Significant differences in time to discharge from hospital were identified between COVID-19 patients with low and high levels of FSC-low monocytes% (χ^2^=8.68, *P*=0.0016, Figure 5A) and FSC-low/FSC-high monocytes% (χ^2^=10.00, *P*=0.0032, Figure 5B). The differences were still significant after being adjusted for age and gender by the Cox models for FSC-low monocytes% (Z=-2.82, *P*=0.0051) and FSC-low/FSC-high monocytes% (Z=-2.80, *P*= 0.0048).

**Figure 5.**
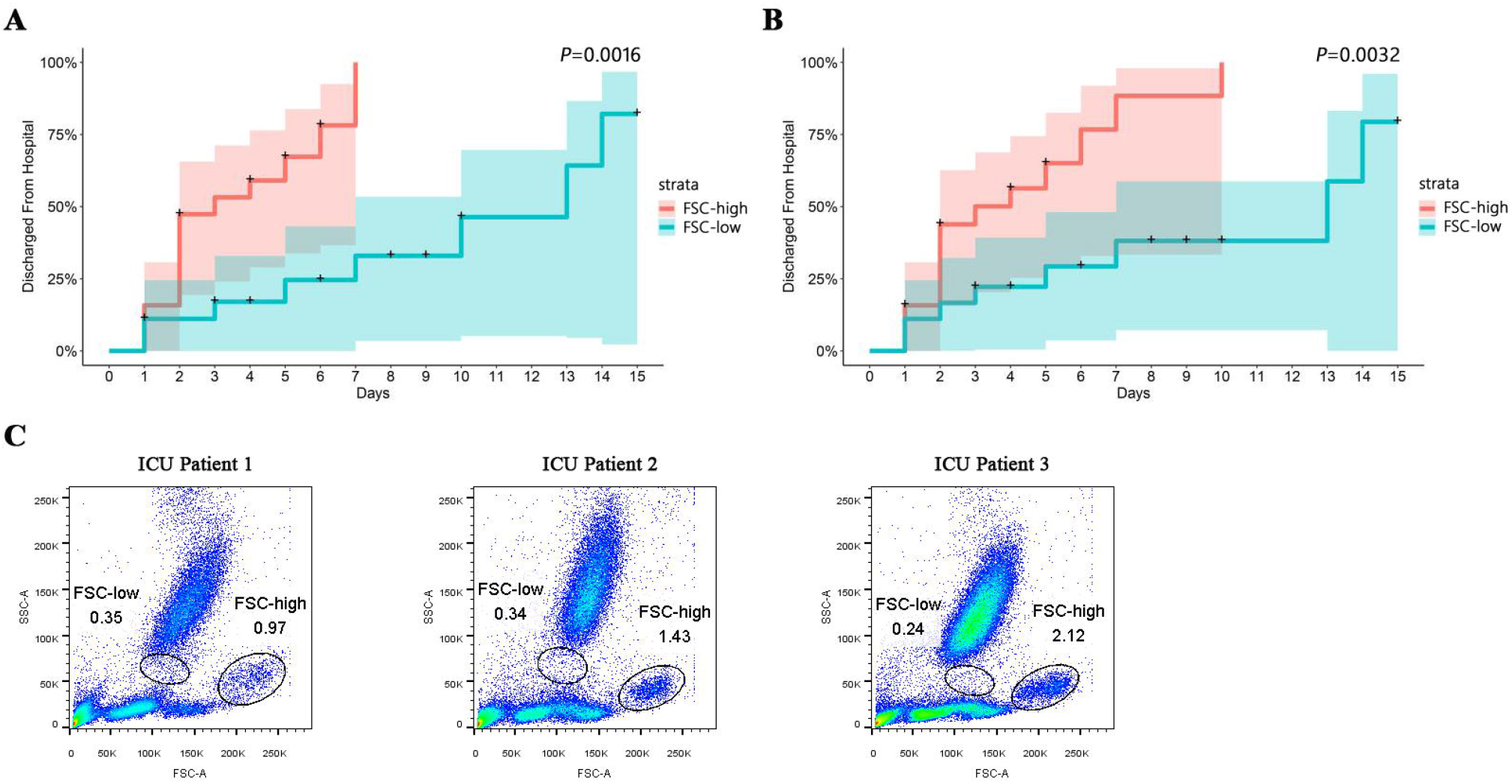
Kaplan-Meier (KM) survival curves obtained from time to discharge from hospital of the COVID-19 patients stratified by levels of FSC-low monocytes and FSC-low/FSC-high monocytes. (A) KM survival curves stratified by levels of FSC-low monocytes. (B) KM survival curves stratified by levels of FSC-low/FSC-high monocytes. (C) The typical FSC/SSC images of the three ICU COVID-19 patients.

### COVID-19 patients admitted to the ICU show a significantly higher ratio of FSC-high population

Of the 28 COVID-19 patients included in this study, three were admitted to the ICU. Focusing on the total number of monocytes and the percentage of FSC-low and FSC-high, we observed an obvious difference from the non-ICU patients. As shown in Figure 5C, all three patients have lower number of monocytes, but a significantly higher percentage of FSC-high than FSC-low population. Moreover, the length of hospitalization of the three ICU patients is more than 45 days, significantly longer that the average 16.24 ± 2.39 days of the non-ICU patients.

### The specific FSC-high population of COVID-19 is not observed in other infectious diseases

To further confirm that FSC-high population is a specific marker for SARS-CoV-2 infection, we investigated whether there is FSC-high population in other infectious diseases. As shown in Figure 6, in 4 cases of HIV, 1 case of HIV with concurrent TB (tuberculosis), 1 case of influenza A virus subtype H1N1, 1 case of Hantaan orthohantavirus (HTNV), and 1 case of Malaria, there was no evidence of a smiliar FSC-high population in these viral and non-viral infections.

**Figure 6.**
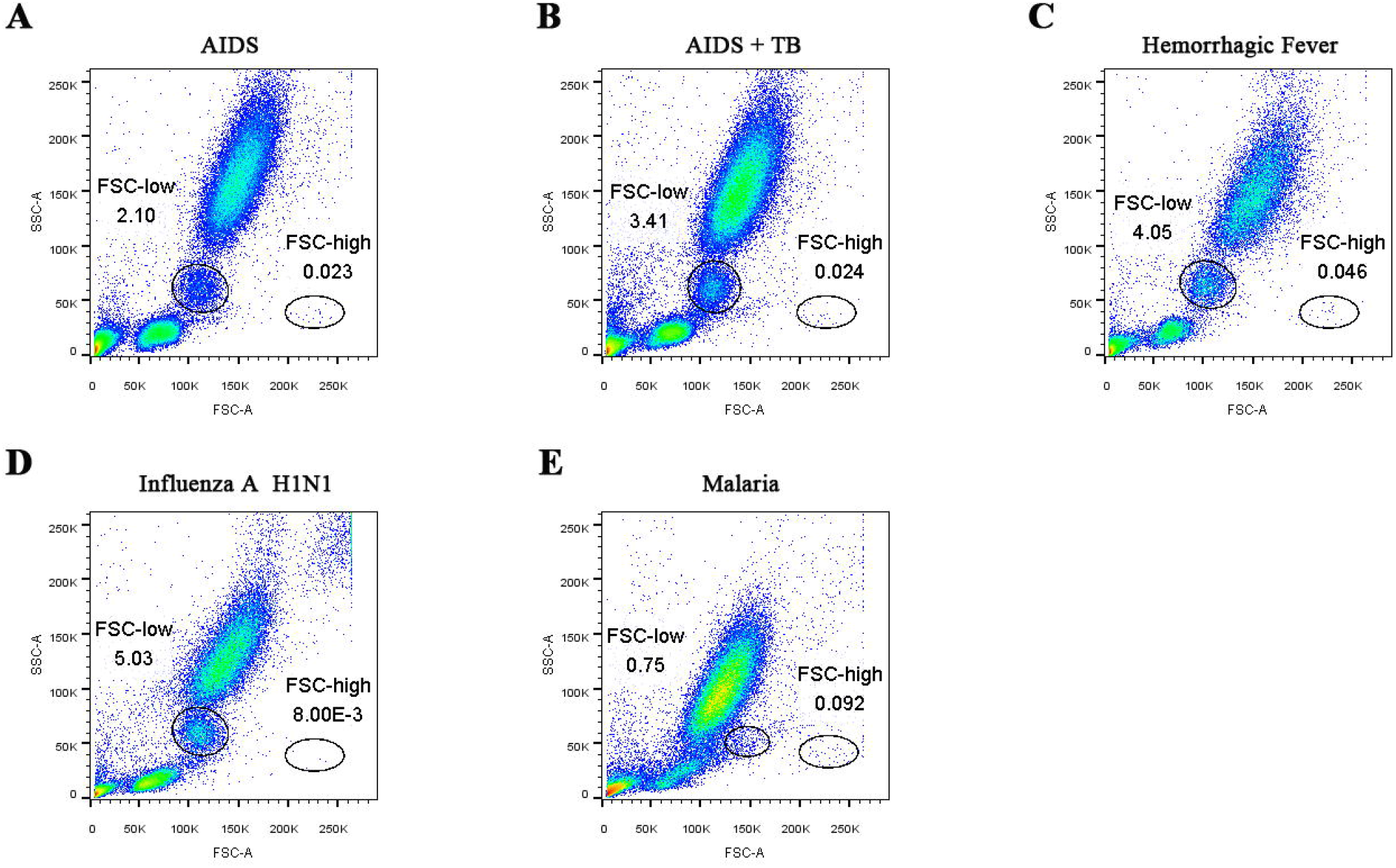
The FSC/SSC images of the viral and non-viral infections. (A) One representative image of 4 HIV patients. (B) AIDS with concurrent TB. (C) Hantaan virus. (D) HIN1. (E) Malaria.

## Discussion

Here we report our findings in 28 patients with confirmed COVID-19 infection with varying degrees of severity. We identified a distinct change in the morphology and function of monocytes/macrophages that was predictive of severity of disease, likelihood of ICU admission, length of hospital stay and full recovery. These monocytes were characterized by a shift in forward scatter on flow cytometry as well as a relative increase in intermediate and non-classical monocytes. These FSC-high monocytes are CD14^+^CD16^+^ and express macrophage markers CD68^+^CD80^+^CD163^+^CD206^+^, which represent an inflammatory monocyte subset not typically seen in healthy controls.

In a review of the causes and consequences of cytokine storm and immunopathology in patients with human corona virus infections that predated the identification of COVID-19, Channappanavar and Perlman identified the following as causes of an exuberant inflammatory response: rapid virus replication, hCoV infection of airway and/or alveolar epithelial cells, delayed type I Interferon (IFN) responses and monocyte/macrophage and neutrophil accumulation.^11^ They highlighted that both SARS-CoV and MERS-CoV encode multiple proteins that antagonize IFN responses and that an early antagonism of the IFN response might delay or evade the innate immune response.^11,12^ They concluded that delayed IFN signaling could further orchestrate inflammatory monocyte/macrophage responses and sensitize T cells to apoptosis resulting in a further dysregulated inflammatory response.^12^ These inflammatory macrophages accumulate in the lungs and are the likely source of pro-inflammatory cytokines and chemokines associated with fatal disease induced by human coronavirus infections, such as SARS and COVID-19. Autopsy findings from patients with COVID-19 mirror these findings.^13^ Interestingly, as we confirmed, monocytes in infected patients are ACE2 positive. Since ACE2 is the receptor used by COVID-19 to gain entry to cells, this suggests the possibility that monocytes in patients with COVID-19 may be directly infected by COVID-19, leading to abortive replication and a delayed type I IFN response in these cells.^12^ Indeed, the SARS-CoV virus has been found to infect monocytes, though replication was poor.^14^ Infection of monocytes by SARS-CoV was associated with inflammatory activation and alterations in immune function-related gene expression.^15,16^ Inflammatory cytokines such as IFN-α, GM-CSF, IL-6 or TNF-α can also stimulate the differentiation of monocytes to macrophages. We have found that treatment high doses of IL-6 or TNF-α are capable of inducing macrophage differentiation in normal peripheral blood samples within 2 days, whereas GM-CSF plus IFN-α requires 5-7 days (data not shown).

Similar to our findings, Zhou and colleagues recently reported on the presence of a significantly higher percentage of CD14^+^CD16^+^ inflammatory monocytes in the peripheral blood of COVID-19 patients compared to normal healthy controls.^17,18^ They also reported that the percentage of CD14^+^CD16^+^ monocytes was much higher in severe pulmonary syndrome patients from ICU. They also showed that these monocytes had the capability of secreting GM-CSF and similar to our findings could also secrete IL-6, which was higher in ICU patients, in association with cytokine storm. They described a model whereby an abnormal TH1 response triggers GM-CSF induced monocyte/macrophage activation leading to an increase in IL-6 producing CD14^+^ CD16^+^ monocytes, which migrate to the lungs and induce subsequent lung damage along with an inflammatory cytokine storm. This is also supported by recent data relating to single cell RNA sequencing of immune cells from bronchoalveolar lavage samples of patients with COVID-19, which provide important insights into the immune microenvironment of COVID-19 infected lungs.^19^ These data show that Ficolin 1 expressing monocyte-derived macrophages, supplant fatty acid binding protein-4 (FABP4) expressing alveolar macrophages as the predominant macrophage subset in the lungs of patients with ARDS. These cells are highly inflammatory and enormous chemokine producers implicated in cytokine storm. Based on these data, a pilot trial of Tociluzumab, an IL-6 receptor antibody approved for treatment of Rheumatoid Arthritis as well as CAR-T associated cytokine release syndrome was recently conducted in a small number of severe COVID-19 patients with promising results.^20-22^ Other treatments for cytokine storm, such as the interleukin-1 (IL-1) receptor antagonist Anakinra and the Janus kinase (JAK) inhibitors may also prove to be useful, though to date there is little or no clinical experience with these agents in treating COVID-19.^23-25^

Given the central role that monocytes appear to play in COVID-19 infection it is important to recognise the limited information provided by most routine automated blood analyzers. Indeed, the contribution of monocytes to the patient’s pathology may be overlooked as patients with severe disease can be monocytopenic.^5^ This may well reflect migration of the inflammatory monocyte/macrophages into the lungs and other affected organs. While morphological examination of peripheral blood films revealed somewhat larger, atypical, vacuolated monocytes, these findings are not very specific. We have shown that simple assessment of FSC by flow cytometry in the context of COVID-19 infection can rapidly identify those patients with an increasing proportion of large, activated, IL-6 and TNF secreting monocytes, who have severe disease and are at greatest risk of ICU admission. In contrast, patients with a high proportion of normal monocytes have better prognosis with earlier recovery and discharge from hospital. These findings appear to be relatively specific for COVID-19 as we have not seen a similar pattern in patients with other viral illnesses, such as H1N1 influenza, HIV, or Hantaanvirus.

In conclusion, in patients with severe COVID-19 infection, monocyte activation and the associated inflammatory response is associated with characteristic changes that can be rapidly identified by a simple blood-based flow cytometry-based assay, and serially monitored. While we acknowledge the limitations of our study, given the small sample size, we feel nevertheless that our findings could be of great help in guiding prognostication and treatment of patients with COVID-19 and merit further evaluation and confirmation in future studies.

## Data Availability

The data used to support the findings of this study are available from the corresponding author upon request.

